# Cross-sectional cycle threshold values reflect epidemic dynamics of COVID-19 in Madagascar

**DOI:** 10.1101/2021.07.06.21259473

**Authors:** Soa Fy Andriamandimby, Cara E. Brook, Norosoa Razanazatovo, Jean-Marius Rakotondramanga, Fidisoa Rasambainarivo, Vaomalala Raharimanga, Iony Manitra Razanajatovo, Reziky Mangahasimbola, Richter Razafindratsimandresy, Santatra Randrianarisoa, Barivola Bernardson, Joelinotahiana Hasina Rabarison, Mirella Randrianarisoa, Frédérick Stanley Nasolo, Roger Mario Rabetombosoa, Rindra Randremanana, Jean-Michel Héraud, Philippe Dussart

## Abstract

As the national reference laboratory for febrile illness in Madagascar, we processed samples from the first epidemic wave of COVID-19, between March and September 2020. We fit generalized additive models to cycle threshold (C_t_) value data from our RT-qPCR platform, demonstrating a peak in high viral load, low-C_t_ value infections temporally coincident with peak epidemic growth rates estimated in real time from publicly-reported incidence data and retrospectively from our own laboratory testing data across three administrative regions. We additionally demonstrate a statistically significant effect of duration of time since infection onset on C_t_ value, suggesting that C_t_ value can be used as a biomarker of the stage at which an individual is sampled in the course of an infection trajectory. As an extension, the population-level C_t_ distribution at a given timepoint can be used to estimate population-level epidemiological dynamics. We illustrate this concept by adopting a recently-developed, nested modeling approach, embedding a within-host viral kinetics model within a population-level Susceptible-Exposed-Infectious-Recovered (SEIR) framework, to mechanistically estimate epidemic growth rates from cross-sectional C_t_ distributions across three regions in Madagascar. We find that C_t_-derived epidemic growth estimates slightly precede those derived from incidence data across the first epidemic wave, suggesting delays in surveillance and case reporting. Our findings indicate that public reporting of C_t_ values could offer an important resource for epidemiological inference in low surveillance settings, enabling forecasts of impending incidence peaks in regions with limited case reporting.

## Introduction

Madagascar reported its first case of coronavirus disease 2019 (COVID-19) on 19 March 2020, in part with a government-sponsored surveillance platform testing all incoming international travelers [1]. Subsequent to this introduction, the first wave of the COVID-19 epidemic was geographically staggered, with early cases in May 2020 largely concentrated in the eastern city of Toamasina, part of the Atsinanana administrative region, followed by a more severe outbreak which peaked in July 2020 in the capital city of Antananarivo, part of the Analamanga administrative region. Test positive rates exceeded 50% at the epidemic peak for both regions and at the national level, indicating widespread underreporting [2], a common feature of COVID-19, for which some 20-40% of infections are thought to be entirely asymptomatic [3–6]. Early reporting on the first epidemic wave in Madagascar indicated an extremely high (56.6%) proportion of asymptomatic cases, based on targeted surveillance of symptomatic patients and their contacts [1].

Madagascar closed its borders to international air travel on 20 March 2020 and, subsequent to identification of the first case, implemented several non-pharmaceutical interventions aimed at curbing epidemic spread, including non-essential business closures, curfews, stay-at-home orders, and mandates for social distancing. These restrictions were relaxed after the first epidemic subsided in September 2020 but have since been re-implemented in the face of a second epidemic wave. In other regions of the globe, widespread efforts to estimate the effective reproduction number, *R*_*t*_, for COVID-19 at national, regional, and local levels [7] have been used to inform public health interventions and retrospectively assess their effectiveness [8]: disease transmission rates are increasing at *R*_*t*_ > 1 and decreasing at *R*_*t*_ < 1. Estimation of *R*_*t*_, or its related counterpart, *r*, the epidemic growth rate [9,10], from available case count data is challenged by limitations or variability in surveillance, uncertainty surrounding the shape of disease parameter distributions, and delays in reporting [8]. Despite the enormity of these challenges in the limited surveillance settings common to many lower- and middle-income countries (LMICs), real-time estimation of *R*_*t*_ from COVID-19 case-counts has been attempted for most regions of the globe [7] and has been implemented locally in Madagascar [11].

Recent methodological advances have introduced a new resource to the epidemiological toolkit by which to conduct real time estimation of epidemic trajectories [12], one that leverages the often-discarded cycle threshold, or C_t_, value, that is returned as an-inverse log-10 measure of viral load from all RT-qPCR-based platforms [13]. After observing that SARS-CoV-2 viral loads—and, as a consequence RT-qPCR C_t_ values—demonstrate a predictable trajectory following the onset of infection [14–16], Hay et al. 2020 showed that the C_t_ value can be used as a biomarker of time since infection and, consequently, be leveraged to back-calculate infection incidence, in a manner analogous to previous work leveraging serological titer information in other systems [17–19]. Probabilistically, a randomly-selected infection is more likely to be early in its infection trajectory when identified during a growing epidemic and later in its trajectory in a declining epidemic [20,21], and as a consequence, the population-level distribution of C_t_ values for any viral infection is expected to shift across the duration of an epidemic. Indeed, low-C_t_-high-viral-load infections have been observed to coincide with growing COVID-19 epidemics and high-C_t_-low-viral-load infections with declining epidemics in several settings [15,22,23]. Exploiting this phenomenon, Hay et al. 2020 developed a method that embeds a within-host, viral kinetics model in a population-level disease transmission model to derive epidemic trajectories from cross-sectional C_t_ samples. Because this method depends on quantitative information captured in the biological sample itself, rather than the relationship between case count and reporting date, C_t_ value estimation more accurately predicts true epidemic trajectories than traditional incidence estimation in settings with uneven surveillance [12].

During the early phase of the COVID-19 epidemic in Madagascar, the Virology Unit laboratory (National Influenza Centre) at the Institut Pasteur of Madagascar (IPM) processed the majority of all SARS-CoV-2 testing samples derived from 114 districts across 6 major provinces in the country. Consistent with findings reported elsewhere [15,22,23], we observed a population-level decline in C_t_ values derived from RT-qPCR-testing in our laboratory, coincident with the epidemic peak across the first wave of COVID-19 in Madagascar. We here adopt the methods presented by Hay et al. 2020 to estimate COVID-19 epidemic growth rates at the national level (2018 population ∼26 million [24]) and in two major administrative regions of Madagascar: Atsinanana (east coast of Madagascar; 2018 population ∼1.5 million [24]) and Analamanga (including Antananarivo, capital city; 2018 population ∼3.6 million [24]). These two regions comprised the geographic epicenter of the first COVID-19 wave in Madagascar; data from other regions were too sparse for epidemiological inference. We evaluate the robustness of this C_t_-based method in comparison with epidemic growth rates derived from more traditional case-count methods applied to the same regions.

## Materials and Methods

### IPM SARS-CoV-2 C_t_ Data

Methods for collection, transport, and processing of SARS-CoV-2 testing samples at IPM have been previously described [1]. Briefly, nasopharyngeal and oropharyngeal swabs were collected at local administrative hospitals in viral transport medium and transported at 4°C to our laboratory for testing. Between 18 March and 30 September 2020, we conducted 34,563 RT-qPCR tests targeting the E, N, Orf1a/b, or S gene of SARS-CoV-2. These tests were carried out across 20,326 discrete samples (many of which were tested across multiple platforms targeting multiple genes), and 17,499 discrete patients, a subset of whom were tested at multiple timepoints. The majority of tests were conducted on individuals who independently sought testing, due to concerns about exposure or symptom presentation, though a subset of samples were derived from efforts to trace and test contacts of positive travelers in the early stages of the pandemic [1]. The reason each patient sought testing was not recorded in the original data.

Due to a dearth of available reagents in the early stages of the epidemic, our lab used seven different WHO-recommended kits and corresponding protocols [25] to assay infection in these samples [1]: Charity Berlin [26], Hong Kong University [27], Da An gene (Da An Gene Co., Ltd. Sun Yatsen University, Guangzhou, China), LightMix® SarbeCoV E-gene plus EAV control (TIB Biolmol, Berlin, Germany), SarbeCoV TibMolBiol (TIB Biolmol, Berlin, Germany), TaqPath^™^ COVID-19 Combo kit (Life Technologies Ltd, Paisley, UK), and GeneXpert (Cepheid, Sunnyvale, CA, USA).

Some 9,493 of those tests, corresponding to 5,310 individuals, were RT-qPCR positive for SARS-CoV-2 infection based on the cut-off positive value for the test in question (Charity Berlin: <= 38; Hong Kong: <=40; Da An: <= 40; LightMix SarbeCoV/SarbeCoV TibMolBiol <= 38; TaqPath <= 37 for 2 of 3 targets; GeneXpert = <= 40). All analyses presented in this paper are derived from these positive test results, as C_t_-values were not reliably recorded following negative results. We further subset our data as appropriate for each analysis of interest.

### Estimating growth rates from IPM case data

We first sought to obtain an estimate of new daily cases reported from our laboratory to the Malagasy government between 18 March and 30 September 2020. To this end, we reduced our dataset to include only sampling from the first reported positive test date for each unique patient; we assumed that reinfection was unlikely within the short duration of our study and that any subsequent positive tests were reflective of longer-duration infections in repeatedly sampled individuals. A patient was considered “positive” for SARS-CoV-2 infection if any test for any SARS-CoV-2 target was positive, and the results of the other samples were not inconsistent with this finding. We then summed cases by date at the national level and for two administrative regions (Atsinanana and Analamanga) that reported the majority of total cases across the study period overall. In total, 5,276 cases were reported from our laboratory across the study period at the national level, 3,505 in Analamanga region and 758 in Atsinanana. Daily cases for the two target regions and for the nation at large are summarized in Fig. 1.

**Fig. 1.**
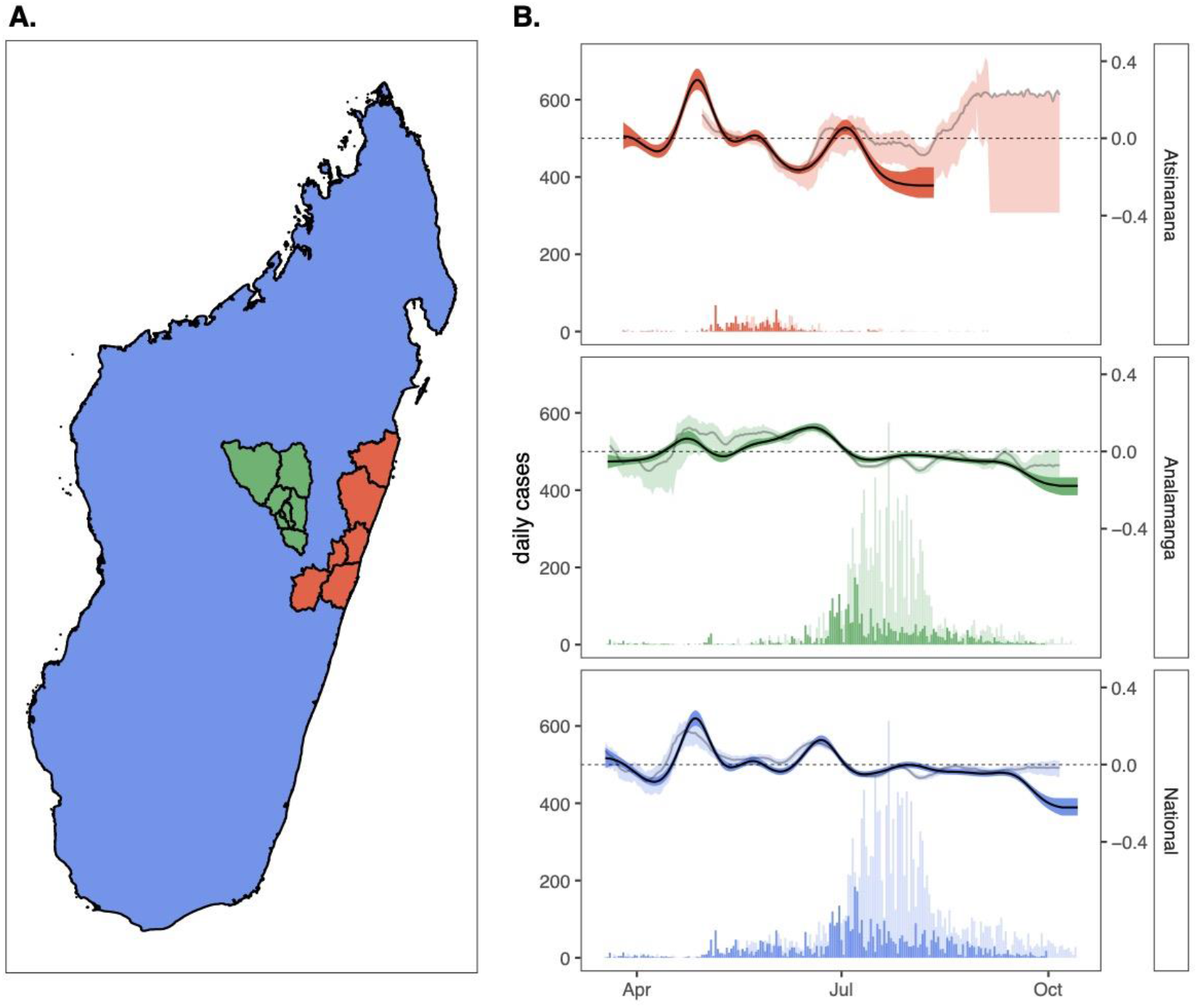
Epidemic growth rate estimates from case count data across the first wave of COVID-19 in Madagascar. **(A.)** Map of Madagascar, colored by regions of case count tabulation, showing the Atsinanana region (orange), the Analamanga region (green), and the National region (blue); note that data analyzed at the National level includes data from both Atsinanana and Analamanga regions, as well as the rest of Madagascar. **(B.)** Time series of new case incidence (lefthand y-axis) across the first wave of COVID-19 in Madagascar (18 March – 30 September 2020), across three focal regions. Darker shading shows data derived from the IPM RT-qPCR platform, while lighter shading depicts data nationally reported and consolidated on [11]. Righthand y-axis shows corresponding epidemic growth rate computed from case count data in EpiNow2 [28], with darker line corresponding to computation from IPM data and lighter line to computation from publicly reported data; background shading around each line depicts the corresponding 50% quantile by EpiNow2 [28].

We applied the opensource R-package EpiNow2 [28] to the daily incidence data to estimate the epidemic growth rate for COVID-19 across the study period. EpiNow2 builds on previous *R*_*t*_ estimation packages [29], using a non-stationary Gaussian process model to estimate the instantaneous time-varying reproduction number, *R*_*t*_, and the corresponding time-varying epidemic growth rate, *r*, while incorporating uncertainty in the generation interval. Following best recommended practices [8], we modeled the SARS-CoV-2 incubation period as a log-normal distribution with a mean of 1.621 days (sd=0.064) and a standard deviation of 0.418 days (sd=0.061) [30] and the generation time interval as a gamma distribution with a mean of 3.635 (sd=0.71) and a standard deviation of 3.075 (0.77) [31]. Since the IPM testing data reported the actual date of sample collection, no reporting delay was incorporated in our growth rate estimation.

### Epidemic trajectories from publicly reported data

To compare our laboratory-derived epidemic growth estimates with those undertaken in real time in Madagascar, we collaborated with colleagues who recorded data on the number of new PCR-confirmed cases reported daily on national television by the Ministry of Health of the Government of Madagascar across the duration of the first epidemic wave. From these daily case estimates, we used the EpiNow2 package [28] to again estimate the epidemic growth rate across the same study period, assuming the same incubation period and general time interval referenced above [30,31]. For these estimates, we followed methods outlined in [32], to additionally model a reporting delay from a log-normal distribution fit to 100 subsamples with 1000 bootstraps from a publicly available linelist that collates data globally for COVID-19 cases for which both infection onset and notification dates are available [33].

### Standardizing C_t_ values across tests and targets

In our next series of analyses, we leveraged information captured in the individual C_t_ value returned from each positive test. To control for extensive variation in qPCR test and target (each of which reported varying thresholds for positivity), we carried out *in vitro* experiments using SARS-CoV-2 isolates from infected patients reporting similar C_t_ values on the TaqPath platform at the time of sampling. Briefly, three SARS-CoV-2 isolates (designated hCoV-19/Madagascar/IPM-00754/2021, hCoV-19/Madagascar/IPM-01263/2021 and hCoV-19/Madagascar/IPM-01315/2021) were obtained and cultured in Vero cells as previously described [34]. Upon infection with SARS-CoV-2, the culture medium was replaced by infection medium containing DMEM, 5 % FBS, antibiotics, 2.5 μg/ml Amphotericin B (Gibco) and 16 μg/ml TPCK-trypsin (Gibco). Virus-containing supernatants, as determined by the presence of cytopathic effect (CPE), were harvested 7 days after infection by centrifugation at 1500 r.p.m. for 10 min. RNA was subsequently extracted from supernatant and subjected to serial dilutions and subsequent testing on six of the seven RT-qPCR platforms used in our population-level dataset (LightMix SarbeCoV and SarbeCoV TibMolBiol were considered equivalent and tested only using the current version of the kit: SarbeCoV TibMolBiol). We fit linear mixed effect regression models in the lme4 [35] package in R to the resulting C_t_ curves returned from each testing platform across the dilution series and used the fitted slope and y-intercept of each regression equation to reproject all Ct values in our dataset to correspond to results returned from the TaqPath N gene test. We report, analyze, and visualize these TaqPath N corrected Ct values in all analyses.

### Generalized additive modeling of the longitudinal C_t_ distribution by region

After observing a population-level dip in the average C_t_ value recovered from our testing platform, roughly coincident with the epidemic peak in the three regions of interest, we asked the broad question, *what is the population level time-trend of SARS-CoV-2 C*_*t*_ *values across these three regions?* To address this question, we compiled all positive tests from the first date of positive testing for each patient, recording the date, region, test, and target that corresponded to each corrected C_t_ value, in addition to the numerical ID and the symptom status (asymptomatic, symptomatic, or unknown) of the patient from which it was derived. Symptom statuses were recorded by medical staff at the timepoint of sampling and merely indicated whether or not the patient presented with symptoms; thus, ‘asymptomatic’ classification did exclude the possibility that the same patient reported symptoms at later or earlier timepoints across the course of infection. The resulting data consisted of 8,055 discrete C_t_ values, corresponding to 5,280 patients, most of whom were tested using multiple tests and/or gene targets of interest. C_t_ values for these positive test results ranged from 6.36 to 39.91. When reprojected to TaqPath N levels, the range shifted from 7.82 to 39.99, such that 507 C_t_ values classed as “positive” by the cutoff thresholds on other platforms exceeded the C_t_ <=37 threshold for positivity on the TaqPath platform. These samples were nonetheless retained for generalized additive modeling (GAM) of longitudinal C_t_ trends but GAM-projected C_t_ values still exceeding the TaqPath cutoff were later excluded in mechanistic modeling of transmission trends fitted to positive data.

Using the mgcv package [36] in the R statistical program, we next fit a GAM in the gaussian family to the response variable of corrected C_t_ value, incorporating a numerical thinplate smoothing predictor of date, and random effects on the categorical variables of test (Charity Berlin, Hong Kong, Da An, LightMix SarbeCoV, SarbeCoV TibMolBio, TaqPath, or GeneXpert), target (E,N,Orf1a/b, or S), and individual patient ID. We refit the model to three different subsets of the data, encompassing the Atsinanana and Analamanga regions, as well as the entire National data as a whole. We then used the resulting fitted GAMs to simulate population-level C_t_ distributions for each date in our dataset, excluding the effects of test and target in the predict.gam function from mgcv. This produced a test- and target-controlled average C_t_ estimate for each positive patient at the timepoint of sampling. We used these GAM-simulated C_t_ distributions to carry out mechanistic model fitting in subsequent analyses, as described below, excluding 15 patients with Ct projections >37, which exceeded the positive threshold for the TaqPath N gene assay (our standard).

### Generalized additive modeling of C_t_ value since time of infection onset

To validate observations from the literature which indicate that the viral load and corresponding C_t_ value follow a predictable trajectory after the onset of SARS-CoV-2 infection [14,15] within our own study system, we next concentrated analyses on a subset of 4,822 Ct values (corresponding to 2,842 unique samples derived from 2,404 unique patients), for which the timing of symptom onset was also recorded. For each of these samples, we randomly drew a corresponding incubation time from the literature-derived log-normal distribution above [30] to approximate the timing of infection onset. To answer the question, *how does C*_*t*_ *vary with time since symptom onset?*, we fit a GAM in the gaussian family to the resulting data with a response variable of C_t_ and a numerical thinplate smoothing predictor of days since infection onset, as well as random effects of test, target, and patient ID. After fitting, we used the predict.gam function from the mgcv package, excluding the effects of target and test, to produce a distribution of C_t_ values corresponding to times since symptom onset (one per each unique patient ID). We used these C_t_ trajectories to estimate parameters for the within-host viral kinetics model described in final methods section below.

### Generalized additive modeling of the relationship between C_t_ value and symptom status

We next asked the question, *does C*_*t*_ *value vary in symptomatic vs. asymptomatic cases?* Our first investigation of this question required only reconsideration of the individual trajectory GAM described above to include additional predictor variables of age and symptom status, in addition to days since infection onset, target, and test. Since symptom status was recorded only at the first timepoint of sampling for each individual, we limited our individual trajectory dataset to a 4,072 datapoint subset of Ct values from 2,404 discrete patients reporting both date of symptom/infection onset and symptom status at the timepoint of sampling; as mentioned previously, ‘asymptomatic’ classification in our dataset included patients reporting symptoms from earlier or later timepoints prior to or following the sampling date. Thus, this GAM tested whether symptom status and C_t_ value interacted merely as a function of the timing since symptom onset (e.g. high C_t_ values were recovered from patients either very early or late in their infection trajectory), or whether independent interactions between symptomatic vs. asymptomatic infections and C_t_ were also present, while also controlling for age.

After observing results, we extended this analysis by applying another GAM in the gaussian family to a 7,937 datapoint subset of the data used to model longitudinal C_t_ trajectories at the National level, which additionally reported symptom status (symptomatic vs. asymptomatic) at the timepoint of first sampling for 5,202 unique patients. Corrected C_t_ values in this data subset ranged between 7.82 and 39.99. This GAM incorporated a response variable of C_t_ and random effects predictor smoothing terms of symptom status, test, target, and patient ID, as well as a numerical smoothing predictor for age of the infected patient.

### Estimating epidemic growth rates from cross-sectional C_t_ values

Finally, following newly-developed methods [12], we sought to estimate the epidemic growth rate across our three regions of interest using cross-sectional C_t_ distributions and compare these results against estimates derived from case count methods described above. To this end, we first fit the within-host viral kinetics model described in Hay et al. 2020 to the test- and target-controlled C_t_ values produced from the above GAM describing C_t_ as a function of time since infection. We used the resulting parameter estimates as informed priors (Table S1) which we next incorporated into two population-level SARS-CoV-2 transmission models applied to our time series data across the three Madagascar regions: a compartmental SEIR model and a more flexible Gaussian process model [12]. Beyond the viral kinetics parameters, we adopted less-constrained priors from the original paper [12] for other epidemiological parameters included in both population-level models (Table S1), then re-fit both transmission models in turn to cross-sectional weekly C_t_ distributions derived from the Atsinanana, Analamanga, and National-level datasets. We fit both models to each dataset using an MCMC algorithm derived from lazymcmc R-package [37], as described in the original paper [12], applying the default algorithm to the Gaussian process fit and a parallel tempering algorithm able to accurately parse multimodal posterior distributions to the SEIR fit. Four MCMC chains were run for 500,000 iterations in the case of the Gaussian process model and three MCMC chains for 80,000 iterations each in the case of the SEIR model, then evaluated for convergence via manual inspection of the resulting trace plots and verification that 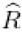, the potential scale reduction factor, had a value <1.1 and the effective population size had a value >200 for all parameters estimated.

After confirming chain convergence, we computed epidemic growth rates from the resulting estimated infection time series and compared results with those derived using more traditional case count methods outlined above. Code and supporting datasets needed to reproduce all analyses are available for download on our opensource GitHub repository at: github.com/carabrook/Mada-Ct-Distribute.

## Results

### Epidemic trajectories from case count data

The first wave of COVID-19 infections in Madagascar, between March and September 2020, was characterized by two subsequent outbreaks: one early, May 2020 peak centered in the eastern port city of Toamasina (region Atsinanana), followed by a second peak in July centered in the capital city of Antananarivo (region Analamanga) (Fig. 1) [1]. Estimation of the epidemic growth rate showed broad agreement in trends at both the national and regional levels, whether computed from IPM testing data assuming perfect reporting of testing date, or from publicly reported national data, including a reporting delay parameterized from a global opensource database (Fig. 1) [33]. Since IPM data comprised just over 30% of nationally reported data throughout the first six months of the Madagascar epidemic, this concurrence in growth rates was unsurprising but nonetheless validates the applicability of the globally parameterized reporting delay for use in Madagascar. In both datasets, we estimated the national level epidemic growth rate to be increasing in the months preceding the two epidemic sub-peaks (in April and in June) and declining beginning in mid-July after the last peak in national case counts (Fig. 1). When IPM data were considered at the regional level, we discovered the April peak to be concentrated in Atsinanana, preceding the Toamasina outbreak and the June peak to be concentrated in Analamanga preceding the Antananarivo outbreak. Growth rate estimation from publicly reported data confirmed this pattern for Analamanga but was not possible for the Atsinanana region due to a lack of clarity in regional reporting.

### Standardizing C_t_ values across tests and targets

All RT-qPCR platforms used in our laboratory demonstrated increases in C_t_ value corresponding to 10-fold dilutions of RNA extracted from the original virus isolate (Table S2), though the estimated slope and y-intercept of each regression varied across the tests and targets considered, with the steepest slope recovered from GeneXpert N-gene tests and the shallowest from the Hong Kong ORF1a/b kits (Fig. S1, Table S3). We used the corresponding slope and y-intercept for each test and platform to transform C_t_ values in all subsequent analyses into values predicted for a TaqPath N-gene platform.

### Longitudinal population-level trends in SARS-CoV-2 C_t_ values across the epidemic wave

We observed a population-level dip in C_t_ values obtained from our SARS-CoV-2 RT-qPCR platform concurrent with the regional peak in cases in May for Atsinanana and June for Analamanga, with both peaks observable in the National data (Fig. 2A). GAMs fit to Atsinanana, Analamanga, and National data subsets explained, respectively, 98.8, 98.9, and 98.9% of the deviation in the data (Table S4). All three GAMs demonstrated statistically significant effects of date, test, and individual patient ID, which contributed to the total deviance capture by each model. GAMs fit to the Analamanga and National data subsets showed an additional significant effect of target on the C_t_ value. Partial effects plots were computed from the resulting GAMs (Fig. S2) following methods described in [38] and demonstrated no significant effects of any particular test or target gene. In general, most variation in C_t_ value beyond that of the individual patient was driven by the significant effect of date across all regions (Table S4).

**Fig. 2.**
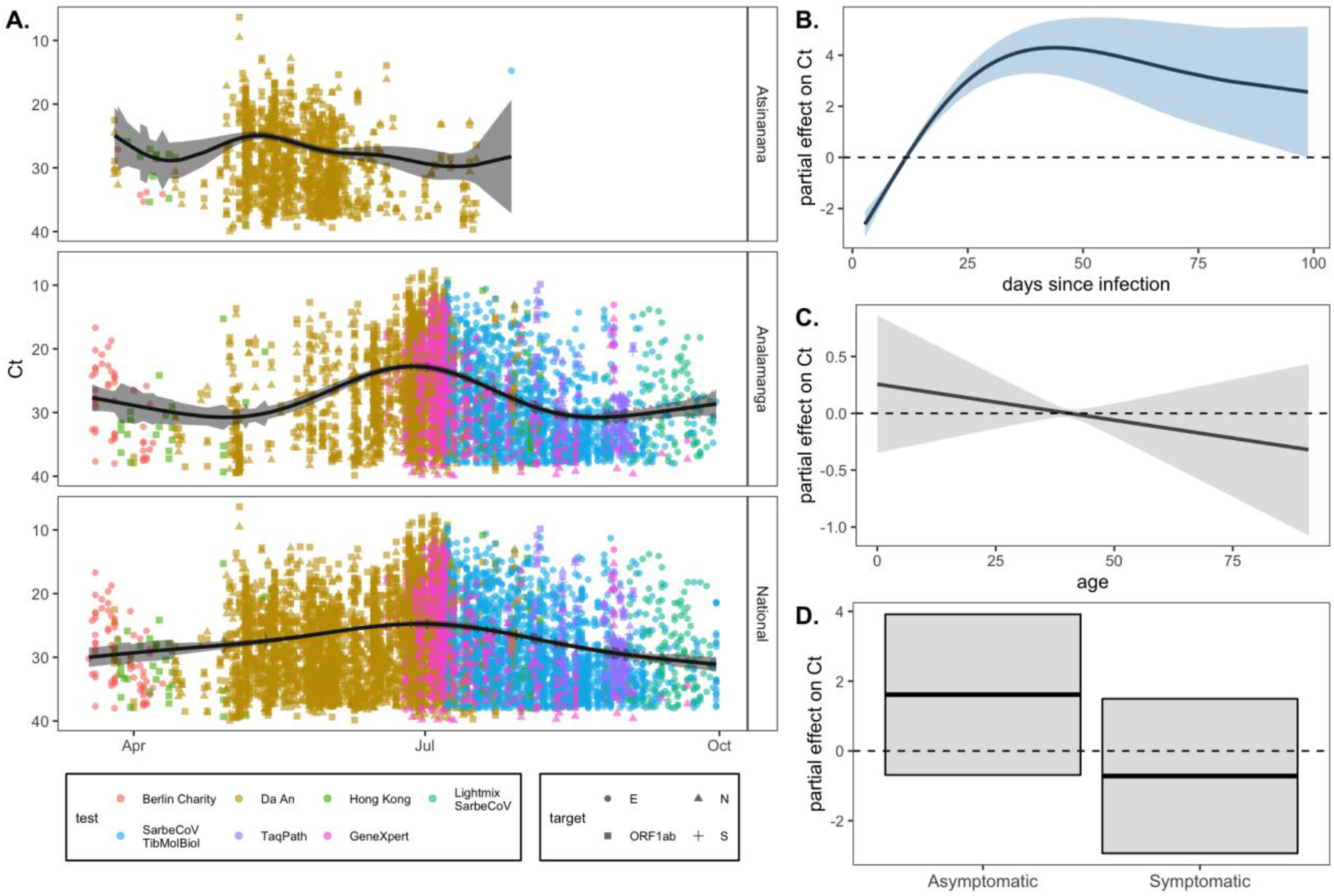
RT-qPCR SARS-CoV-2 C_t_ value as a biomarker of population-level epidemic pace and individual infection trajectory. **(A.)** Population-level SARS-CoV-2 corrected Ct values from IPM RT-qPCR platform across three Madagascar regions from March-September 2020. C_t_ values are colored by the test and shaped by the target from which they were derived (legend), though note that all C_t_ values were first corrected to TaqPath N gene range. Black trend line gives the output from a gaussian GAM fit to these data (Table S4), excluding the effects of target and test, which were also included as predictors in the model; 95% confidence intervals by standard error are shown in translucent shading. Partial effects of each predictor are visualized in Fig. S2. Righthand plots visualize partial effects of **(B.)** days since infection, **(C.)** patient age, and **(D.)** patient symptom status on C_t_ value from our individual trajectory GAM (Table S5). Significant predictors are depicted in light blue and non-significant in gray (Table S5).

### Individual trends in SARS-CoV-2 C_t_ values across the trajectory of infection

The SARS-CoV-2 C_t_ value also demonstrated a predictable trajectory from the timing of onset of infection. Our GAM fit to data reporting a date of symptom onset (which we converted to a date of infection onset) and incorporating a predictor smoothing term of days since infection onset, and random effects of test, target, and patient ID explained 92.7% of the deviance in the data and demonstrated statistically significant effects of all predictor variables, including days since infection onset (Table S5). These findings confirmed that C_t_ value can be used as a biomarker of time since infection, validating the applicability of methods outlined in [12] for our Madagascar data.

### Relationship between symptom status and SARS-CoV-2 C_t_ value

As an extension of the individual trajectory analysis, we hypothesized that C_t_ value would likely be linked to symptom status, since many infection trajectories begin with a brief presymptomatic phase, progress to symptom presentation, then become asymptomatic during recovery [14,15]. The first GAM we employed to address this question considered age and symptom status as additional predictor variables in our individual trajectory analysis. This final GAM explained 98.5% of the deviation in the data and included significant effects of days since infection onset, symptom status, test, target, and patient ID (Table S5). Despite the significance of symptom status as a predictor variable in the GAM overall, partial effects plots demonstrated no significant association between asymptomatic status and high C_t_ values or symptomatic status and low C_t_ values, while controlling for age (Fig. 2B, 2C, 2D). These results suggest that, in our dataset, C_t_ value varies predictably with an individual’s infection trajectory regardless of symptom classification or age of the patient, further validating its adoption as a robust biomarker of time since infection (Table S5).

We additionally extended this analysis to our National-level C_t_ dataset, including a predictor variable of symptom status, in addition to test, target, patient age, and patient ID in longitudinal GAMs. This model explained 98.9% of the deviation witnessed in the data, including significant effects of test, target, patient ID, and symptom status (Table S6). Test and target were here included as control variates only and cannot be considered for prediction, as both co-varied with date, which was not used as a predictor in this model. In this model, partial effects plots indicated a significant association of asymptomatic status with high C_t_ values and symptomatic status with low C_t_ values (Fig. S3), even when controlling for effects of age; as this larger dataset did not report date of symptom/infection onset, it is likely that this association co-varied with the timing of infection onset, suggesting that previous reports of a high proportion of asymptomatic infections in Madagascar [1] could reflect a high proportion of pre- or post-symptomatic infections.

### Epidemiological dynamics inferred from cross-sectional C_t_ distributions

After confirming the predictable pattern of C_t_ value across an individual’s infection trajectory, and the predictable decline in population-level C_t_ in conjunction with the epidemic peak, we used our individual trajectory GAM to simulate a distribution of C_t_ values across a 50-day duration of infection and fit the within-host viral kinetics model described in [12] to the resulting data (Fig. S4). The model demonstrated a good fit to the data, and estimated posterior distributions for viral kinetics parameters were largely on par with those used previously in models of SARS-CoV-2 dynamics in Massachusetts, though the modal C_t_ value at peak viral load was slightly lower in our Madagascar dataset (Fig S4; Table S1).

After fitting the within-host model, we next used longitudinal population-level GAMs (Fig. S2) to generate weekly cross-sectional C_t_ distributions, controlled for test and target, across our three regions of interest. As expected, weekly cross-sectional C_t_ distributions demonstrated a shift across the duration of the epidemic wave; with lower C_t_ values temporally correlated with high growth rates estimated from case count data (Fig. 3).

**Fig. 3.**
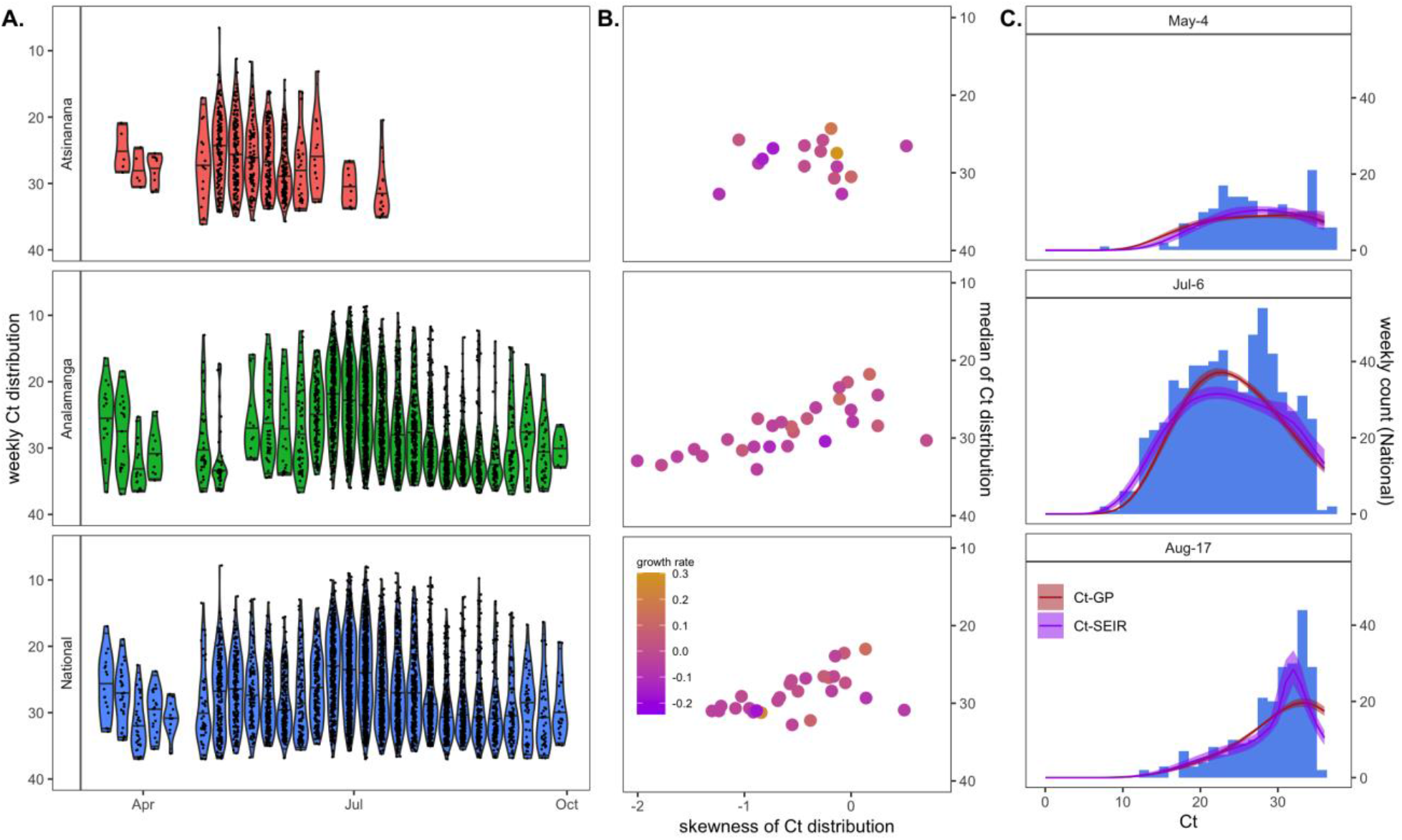
Population-level C_t_ distribution reflects epidemic dynamics of the first wave of COVID-19 across three Madagascar regions. **(A.)** Simulated weekly C_t_ distributions by Madagascar region, derived from population-level longitudinal GAMs (Fig. 2A), excluding random effects of test and target. **(B.)** Higher skew and lower median C_t_ from each cross-sectional Ct distribution in (A.) were loosely associated with higher epidemic growth rates from the corresponding week, here derived from EpiNow2 estimation from IPM case count data (Fig. 1B.) **(C.)** Cross-sectional C_t_ distributions from Analamanga time series in (A.) were fit via Gaussian process (GP) and SEIR mechanistic models incorporating a within-host viral kinetics model. Modeled C_t_ distributions are shown as solid lines (GP=red; SEIR=purple), with 95% quantiles in surrounding sheer shading. Both models effectively recapture the shape of the C_t_ histogram as it changes (skews left) across the duration of the first epidemic wave. Model fits to the full time series of C_t_ histograms across all three regions are visualized in Fig. S7, S8, S9.

Finally, we used the viral kinetics posterior distributions resulting from the within-host viral kinetics model fit as prior inputs into SEIR and Gaussian process population-level epidemiological models, which we fit to the weekly cross-sectional C_t_ data. MCMC chains generated in the fitting process demonstrated good convergence (Fig. S5, Table S7, Table S8) and produced posterior distributions for all parameters on par with those estimated in previous work (Table S1, Fig. S6), which effectively recaptured cross-sectional C_t_ value histograms across the target timeseries in all three regions (Fig. 3, Fig. S7-S9) [12]. From the resulting fitted models, we simulated epidemic incidence curves, which we used to compute growth rate estimates across the duration of the first epidemic wave in each of the three regions (Fig. 4). We compared these estimates to growth rates inferred from case count data; patterns from both SEIR and Gaussian process models were largely complementary, though the more flexible Gaussian process model demonstrated less extreme variation in epidemic growth rate. Both C_t_-model fits demonstrated similar patterns to epidemic trajectories estimated from incidence data, with increasing growth rates in the months preceding both epidemic sub-peaks (April and June) and decreasing growth rates beginning in July. Nonetheless, growth rate estimates derived from the C_t_ model slightly preceded those estimated from case count data. The C_t_ model fits further predicted uncertainty in growth rate directionality towards the end of the study period for the Analamanga and National-level data, while incidence estimation projected decreasing cases at this time. This finding suggests that cross-sectional C_t_ distributions indicated a possible epidemic resurgence which was overlooked by growth rates estimated from declining incidence. If incidence declined in part due to declining surveillance, as was the reality at the end of Madagascar’s first epidemic wave [1], only the C_t_ method remained robust to the possibility of epidemic renewal.

**Fig. 4.**
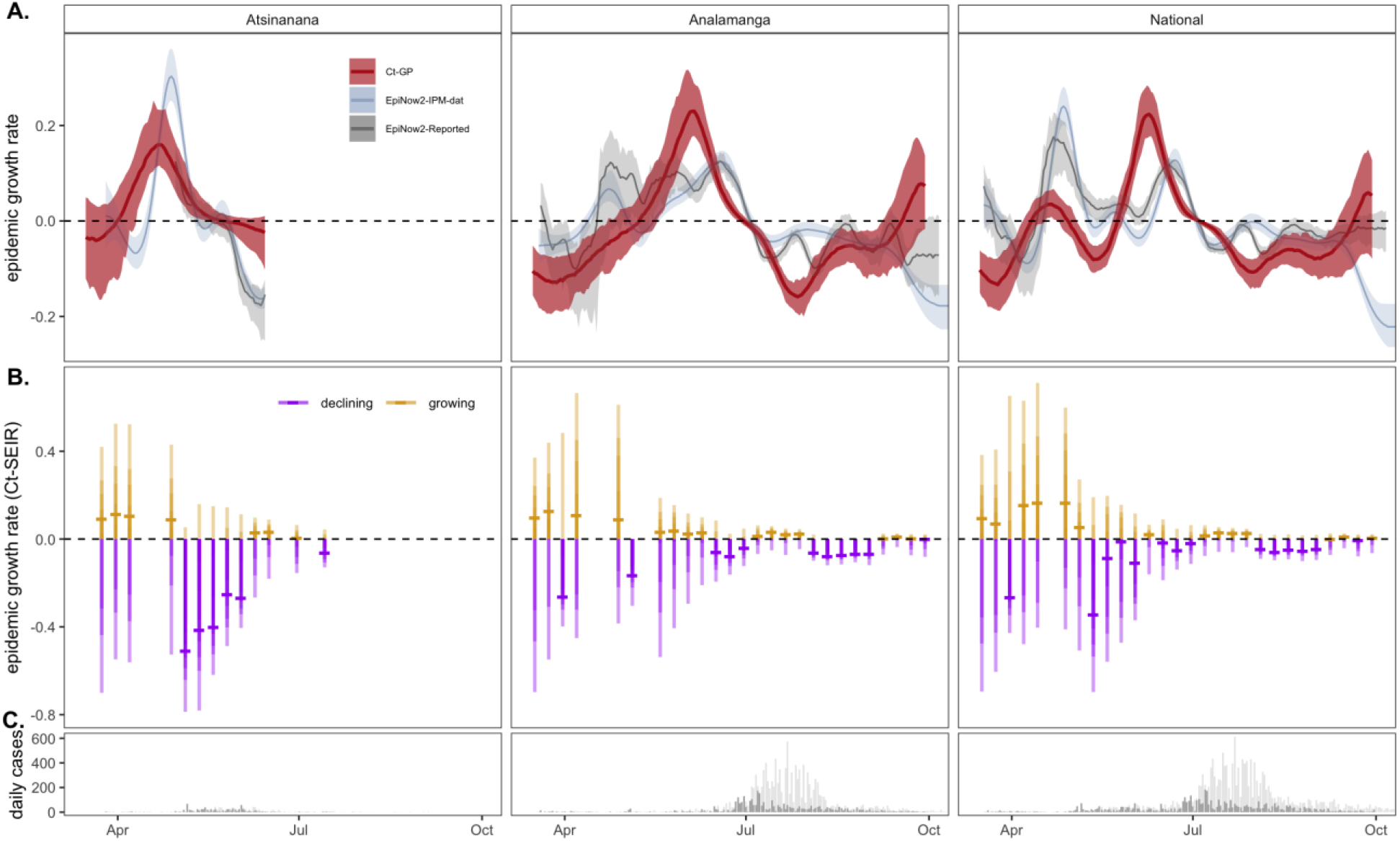
Epidemic growth rate estimates from mechanistic model fits to population-level C_t_ distributions across the first wave of COVID-19 in three Madagascar regions. **(A.)** Comparison of COVID-19 epidemic growth rates from March-September 2020, estimated from IPM (blue) and publicly reported (gray) case count data using EpiNow2 [28] with estimates derived from Gaussian process (GP; red) mechanistic model fit to the time series of C_t_ distributions (Fig. 3A). Median growth rates are shown as solid lines, with 50% quantile on case-based estimates and 95% quantile of the posterior distributions from C_t_-based estimates in corresponding sheer shading. **(B.)** Growth rate estimates from individual SEIR C_t_-model fits to each C_t_-distribution shown in Fig. 3A; median growth rates are given as horizontal dashes, with the 95, 70, 50, and 20% of the posterior distribution indicated by progressively darker coloring. Estimates >0 (indicating growing epidemics) are depicted in gold and <0 (indicating declining epidemics) in purples. **(C.)** Raw case count data from the time series (dark=IPM data; light=publicly reported data) is shown for reference.

## Conclusions

Real-time estimation of epidemiological parameters, including the time-varying effective reproduction number, *R*_*t*_, and the related instantaneous epidemic growth rate, *r*, has played an important role in guiding public health interventions and policies across many epidemic outbreaks, including COVID-19 [39–41]. In Madagascar, an opensource platform [11] was developed shortly after the introduction of COVID-19 in March 2020, to collate and visualize publicly reported data and estimate *R*_*t*_ using traditional methods applied to daily reported incidence [28,29]. We here compare the results from this platform applied to the first epidemic wave in Madagascar, with new estimates of the time-varying epidemic growth rate applied to our own laboratory data across the first epidemic wave—including those derived using a novel method based on the cross-sectional C_t_ value distribution at the time of sampling [12].

We find our new estimates to be largely congruent with those predicted from publicly reported data, demonstrating a pattern of increasing epidemic growth rates prior to a peak in cases, which occurred first in May 2020 in the Atsinanana region, followed by a second outbreak in July 2020 in the Analamanga region. Critically, our growth rate estimates derived using novel methods applied to the C_t_ distribution over time slightly precede those estimated from incidence data. As previous work has demonstrated C_t_ estimation to offer a more robust approximation of true dynamics under limited surveillance scenarios [12], these findings suggest that incidence-based methods to estimate epidemic trajectories in Madagascar may be underestimating the true pace of the epidemic, likely as a result of underreporting. Additionally, C_t_-based methods adopted by a single laboratory allow for estimation of epidemic growth rates even in the absence of publicly reported case counts: in October 2020, the Malagasy Ministry of Health shifted its daily COVID-19 case notifications to weekly, interfering with incidence-based approaches to estimate epidemic trajectories [11]. C_t_-based approaches, instead, should be robust to this variation in reporting, offering a powerful tool for public health efforts in low surveillance settings. Indeed, our analysis demonstrates that C_t_-based epidemic growth rates show uncertain directionality towards the end of the first wave of COVID-19 in Madagascar, presaging eventual epidemic resurgence, while incidence-based rates categorically declined due to both truly declining cases and declining surveillance. Incidence-based growth rate estimation ceased during the continued limited surveillance period from October 2020 through March 2021 [11]; had C_t_-based methods been available at the time, it is possible that the current second wave could have been predicted and mitigated by earlier rollout of public health interventions.

Statistical analysis of our C_t_ data indicates that C_t_ values vary predictably with days since onset of infection, allowing viral kinetics data to be leveraged for population-level estimation of epidemiological patterns. In our system, this pattern held even after controlling for the effects of age and symptom status on the C_t_ trajectory, further validating the applicability of C_t_ value as an indicator of time since infection. Nonetheless, in future work, it mat be possible to fit unique viral kinetics trajectories for different classes of people; for example, older age cohorts or cohorts of people infected with more transmissible variants may be better represented by lower average C_t_ trajectories than the population as a whole [15,42]. Our application of generalized additive models to both individual infection trajectory and population-level C_t_ distributions offers an effective means by which to control for variation in test and target across diverse RT-qPCR platforms to generate C_t_ values for epidemiological inference which represent a reliable average of population-level patterns overall.

We acknowledge the limitations of our current method, especially as it relates to testing biases. During the earliest phases of the epidemic in Madagascar, testing resources were limited in our laboratory, which may have biased sample intake towards high-viral-load, low-C_t_-value cases that could bias epidemiological inference towards increasing growth rates even after the epidemic has, in reality, already begun to decline. As the epidemic ensued, however, the Madagascar Ministry of Health focused sampling on symptomatic patients and their suspected contacts, leading to a high proportion (56.6%) of reported asymptomatic infections in our dataset [1], which may have instead prematurely biased inference towards a declining epidemic. Nonetheless, our Ct-based projections of epidemic trajectories do not appear to underestimate realized trends, suggesting that our method was robust to these inconsistencies.

We apply a novel method leveraging within-host viral load data that is currently largely overlooked in the epidemiological literature to describe the dynamics of the first wave of COVID-19 in Madagascar. Our approach validates an important new tool for epidemiological inference of ongoing epidemics, particularly applicable to limited surveillance settings characteristic of many lower- and middle-income countries. We advocate for public release of real time data describing the C_t_ value distribution, in addition to daily case counts, to improve epidemiological inference to guide public health response and intervention.

## Supporting information

github.com/carabrook/Mada-Ct-Distribute

## Data Availability

All data referred to in the manuscript are available.

## Acknowledgements

This work was supported by the US National Institutes of Health [1R01AI29822-01]; the Bill & Melinda Gates Foundation, Seattle, WA [GCE OPP1211841]; and the Innovative Genomics Institute at UC Berkeley, Berkeley, CA [COVID-19 Rapid Response Grant]. The authors thank Malavika Rajeev, Tanjona Ramiadantsoa, and Benjamin Rice for help in establishing the Madagascar COVID-19 dashboard.

## Supplementary Figures

**Fig. S1.**
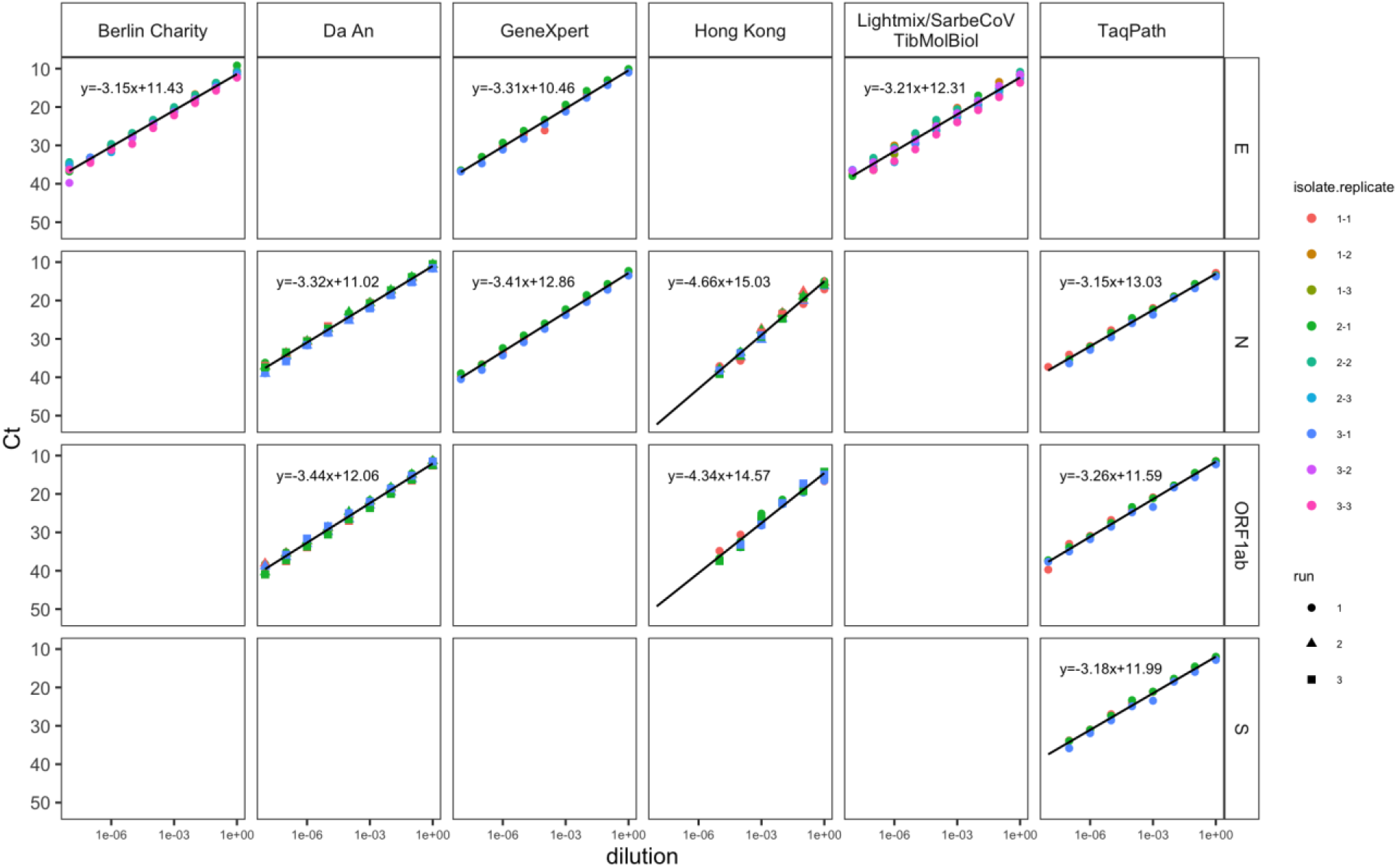
Ct-dilution curves of RT-qPCR tests in tissue culture. RNA isolates from three patients positive with comparable Ct values for SARS-CoV-2 infection underwent serial dilutions (x-axis) RT-qPCR assay (y-axis) across six RT-qPCR platforms used in our laboratory during the first wave of the COVID-19 epidemic (Table S2). We fit linear mixed effect regression models in the lme4 R package [35] to the resulting dilution series across multiple runs and replicates (shapes, colors) for each isolation to establish a trend line (lines and equations). We used the resulting slope and y-intercept (Table S3) to standardize all Ct values in our dataset in terms of TaqPath N-gene assays for downstream analysis.

**Fig. S2.**
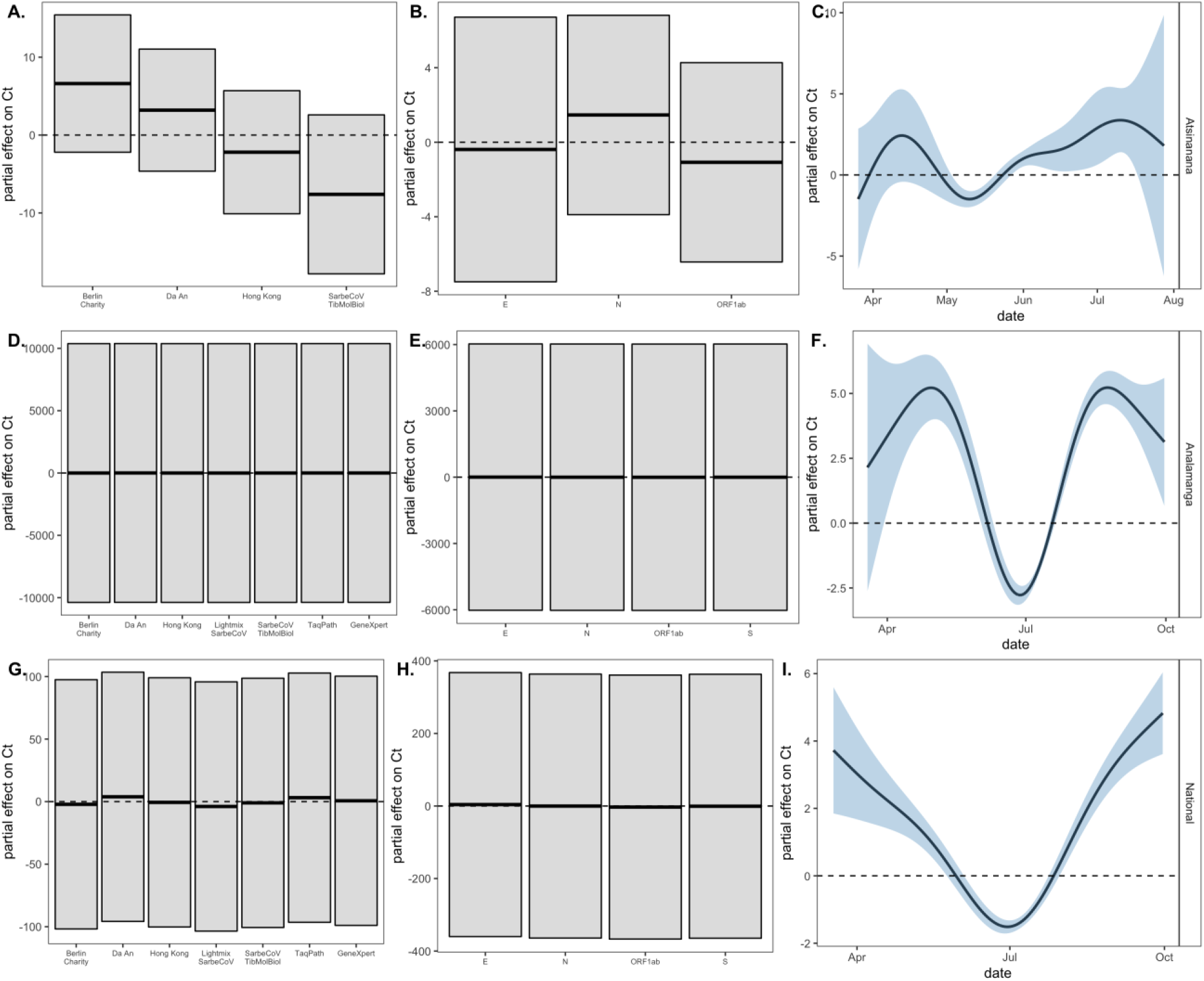
Partial effects of test, target, and date on SARS-CoV-2 Ct values across three Madagascar regions. Partial effects of GAMs fitted to longitudinal C_t_ data from **(A**.,**B**.,**C.)** Atsinanana region, **(D**.,**E**.,**F.)** Analamanga region and **(G**.,**H**.,**I.)** National data. Model outputs are summarized in Table S4 and visualized in Fig. 2A. (main text). Significant partial effects determined by holding all other variables constant (see [38] for methods) are depicted in blue and insignificant partial effects in gray. Columns demonstrated relative effects of RT-qPCR test (A.,D.,G.), target gene (B.,E.,H), and date (C.,F.,I.). Note that patient ID was also included in each model as a (significant) random effect.

**Fig. S3.**
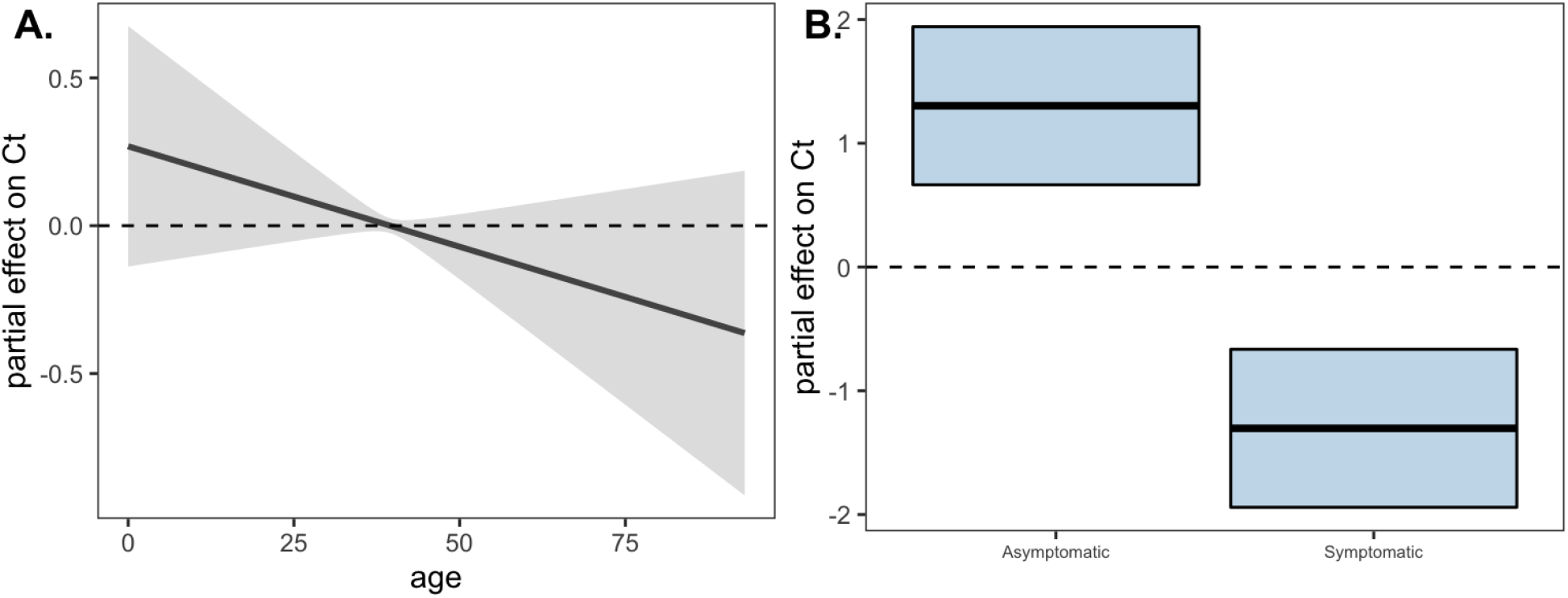
Partial effects of age and symptom status from population-level GAM. Partial effects of GAM fitted to national Ct data with response variable of correct C_t_ and predictor smoothing terms of test, target, and patient ID (as random effect controls) as well as **(A.)** age and **(B.)** symptom status. Model outputs are summarized in Table S6. Significance is indicated by blue shading: the significant effect of symptom status on Ct is likely the result of time since onset of infection (i.e. ‘asymptomatic’ patients were either very early or very late in their infection trajectory), which was not specified in these data.

**Fig. S4.**
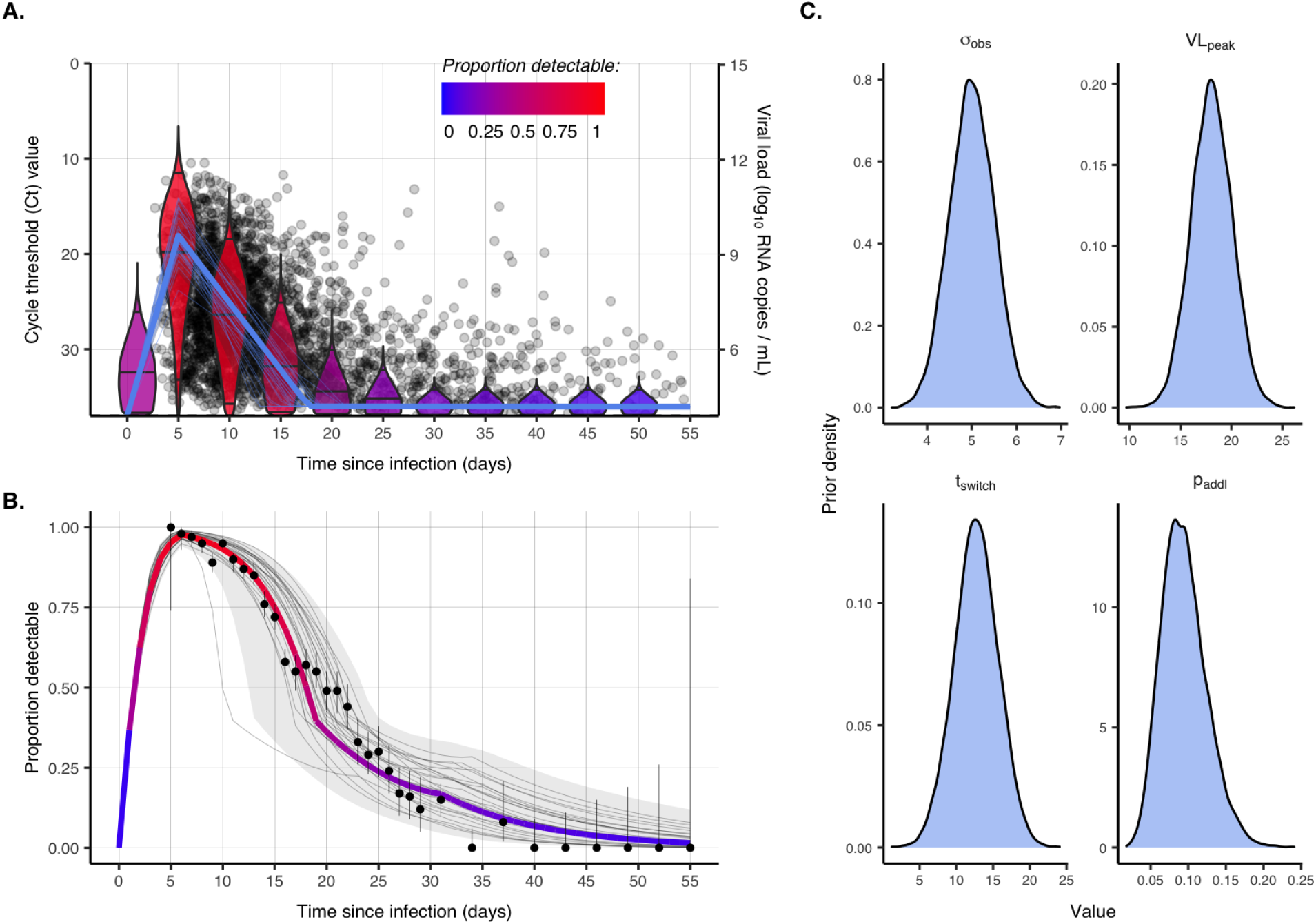
Fitting the within-host viral kinetics model to Madagascar data. Figure recapitulates Fig. S2 from [12] for our Madagascar data. The thick blue line in **(A.)** gives the fitted viral load trajectory for the Madagascar data output from the individual trajectory GAM summarized in Table S5 but excluding the effect of test and PCR-target. Madagascar data are shown as translucent colors in the background. Thin blue lines surrounding the thick line are trajectories from prior draws within the 95% quantile, and violin plots show the distribution of detectable Ct values post infection inferred from the fitted trajectory (blue line). As in [12], violins are colored by the proportion of Ct values above the limit of PCR detection (10^3^ RNA cp/µl). Panel (B.) shows the least-squares based fit (colored line) to the proportion of patients with detectable SARS-CoV-2 in upper respiratory tract samples on each day post symptom onset from Borremans et al. [16]. As in [12], black dots and lines show proportion positive and 95% confidence intervals. Faint grey lines show proportion detectable over time from prior draws, and faint grey ribbon shows 95% quantiles. Panel (C.) gives the assumed prior densities for viral kinetics model parameters fit to Madagascar data (Table S1).

**Fig. S5.**
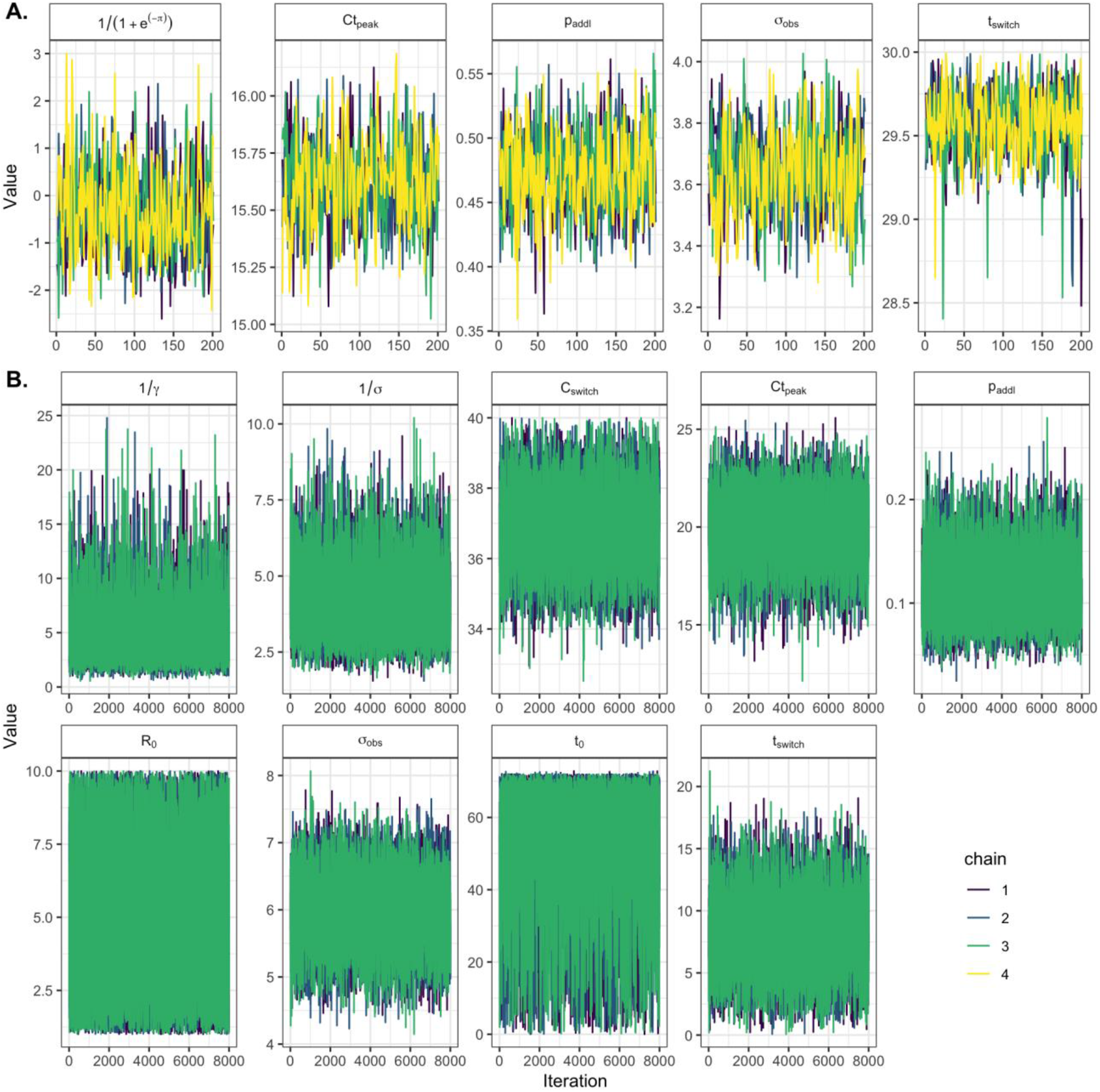
Example trace plots for MCMC-fit of Gaussian process and SEIR Ct-models. Trace plots returned from MCMC fitting of **(A.)** Gaussian process (4-chains, standard MCMC) and **(B.)** mechanistic SEIR models (3 chains, parallel tempering MCMC) to Madagascar National-level time series of Ct distributions. Traces in **(B**.) show results from fitting to timepoint 77, the Ct distribution from the week of April 27, 2020. Parameter details are given in Table S1. All traces show good convergence, but as in [12], we observed that SEIR models demonstrated clear multi-modality in some parameter distributions, chiefly *R*_*0*_ and *t*_*0*_. Data can be explained by either high *R*_*0*_ and low *t*_*0*_ or low *R*_*0*_ and high *t*_*0*_.

**Fig. S6.**
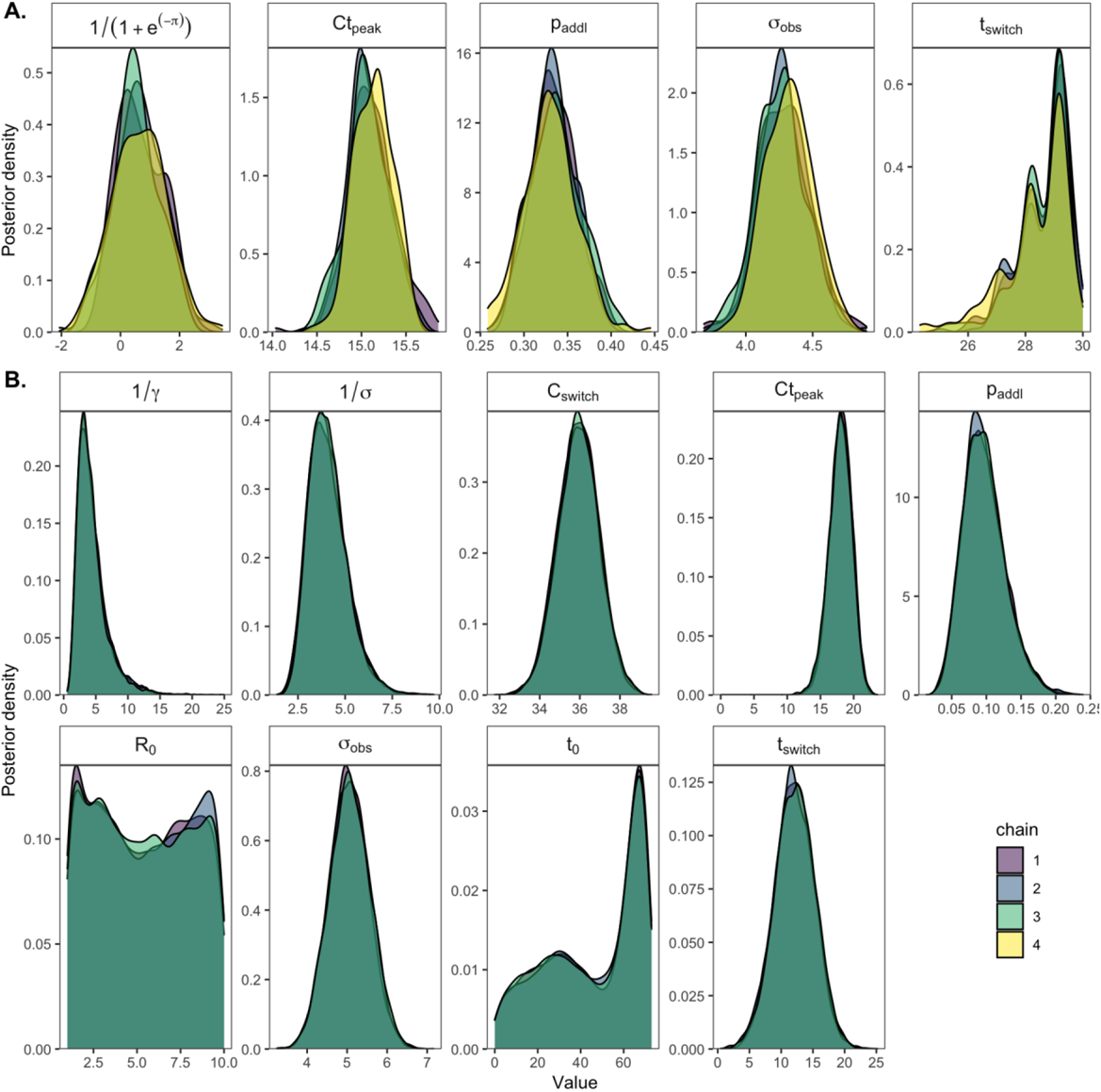
Posterior distributions for fitted SEIR and Gaussian process models to Madagascar Ct timeseries. Posterior distributions for all parameters inferred from MCMC fitting for **(A.)** Gaussian process (4-chains, standard MCMC) and **(B.)** mechanistic SEIR models (3 chains, parallel tempering MCMC) to Madagascar National-level time series of Ct distributions. Parameter details are given in Table S1. As in Fig. S5, distributions in (B.) show results from fitting to timepoint 77, the Ct distribution from the week of April 27, 2020. Multimodality in *R*_*0*_ and *t*_*0*_ is also evident here: data can be explained by either high *R*_*0*_ and low *t*_*0*_ or low *R*_*0*_ and high *t*_*0*_.

**Fig. S7.**
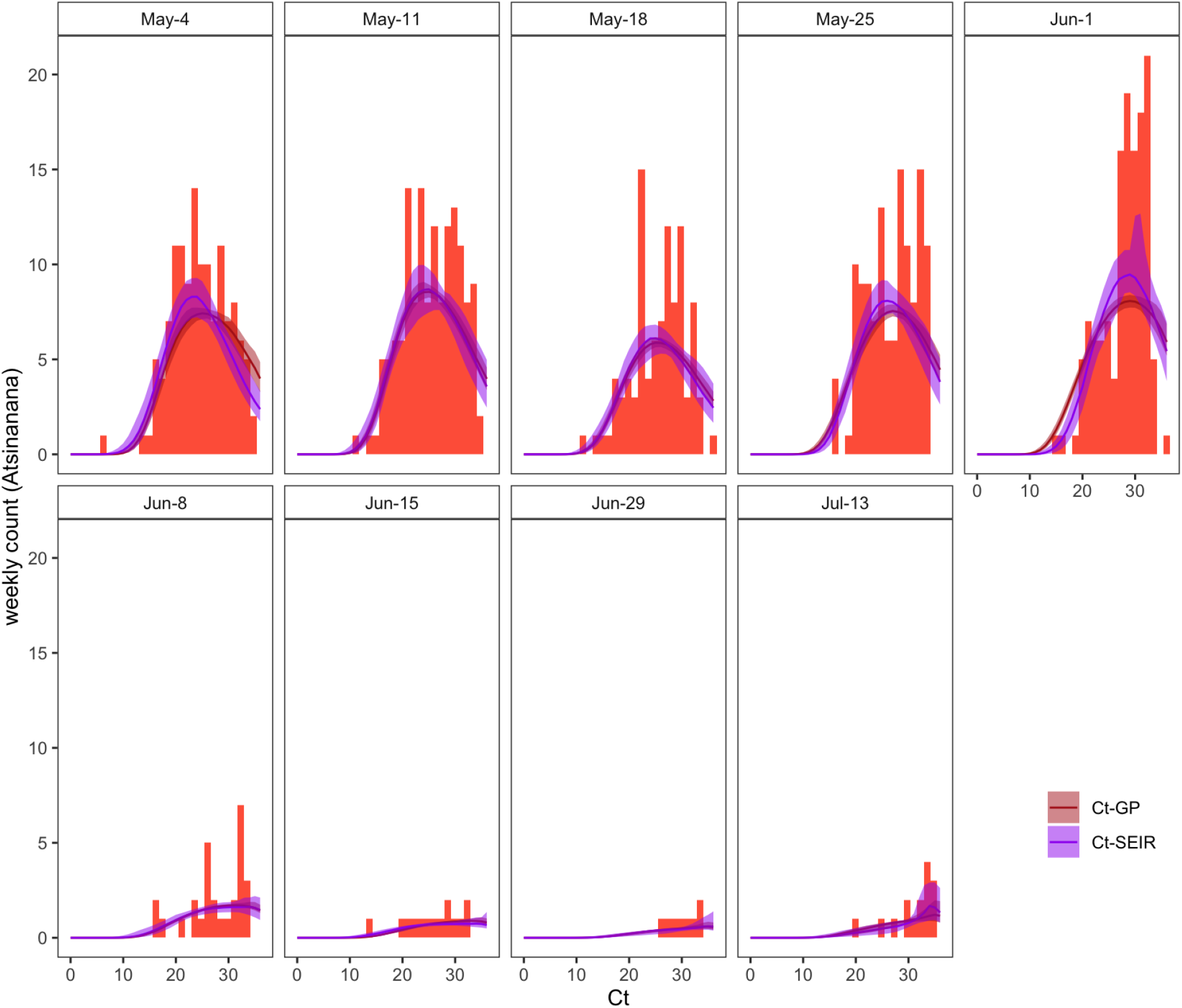
Gaussian process and SEIR Ct-model fits to cross-sectional histograms of Ct values across the Atsinanana time series. Extension of Fig. 3C (main text) depicts the weekly histogram of Ct values from the National timeseries in blue, and the resulting Ct distributions at each timepoint from fitted Gaussian process (red) and SEIR (purple) models. Thick line gives the median of distribution for each fit and translucent shading corresponds to the 95% quantiles.

**Fig. S8.**
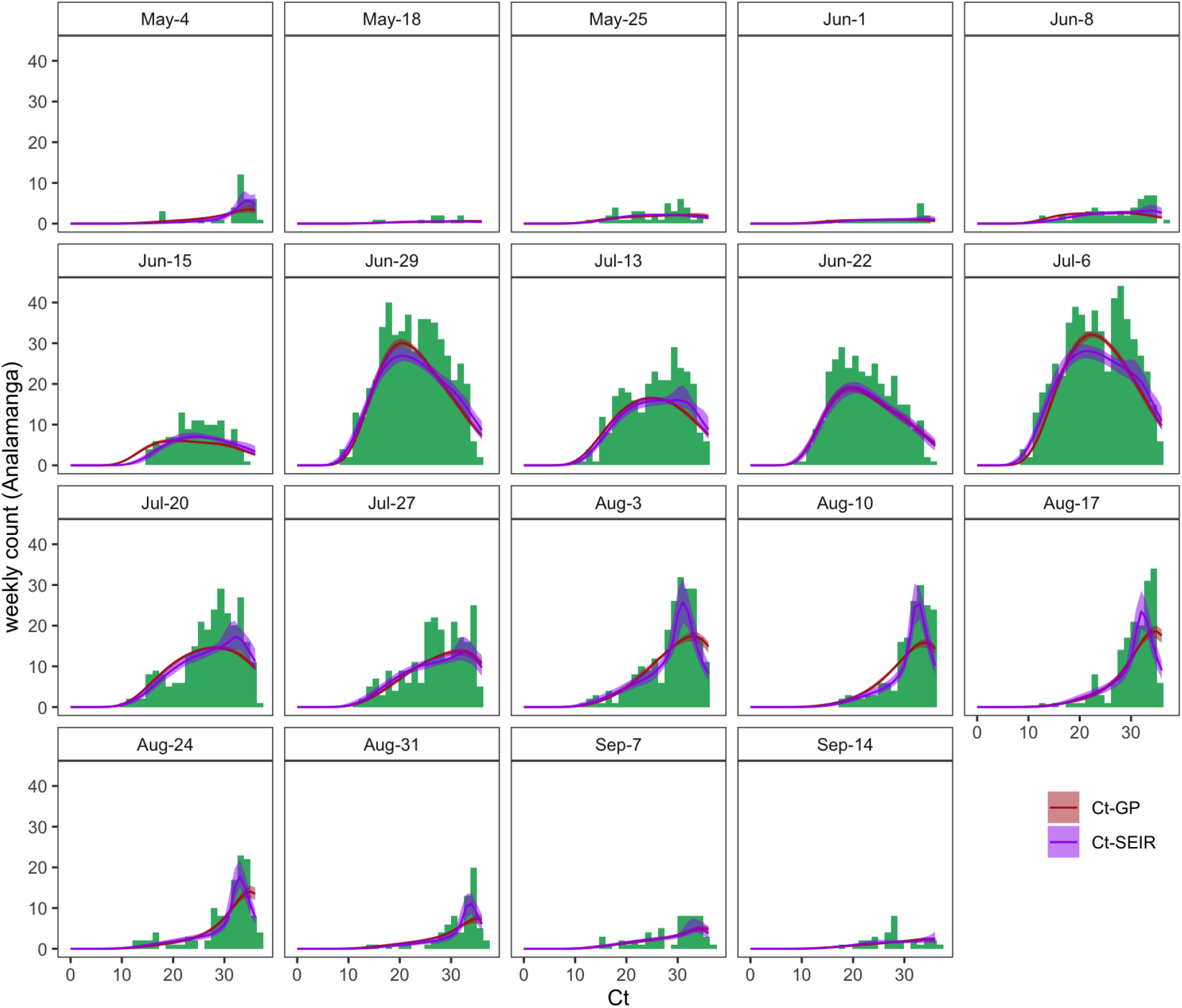
Gaussian process and SEIR Ct-model fits to cross-sectional histograms of Ct values across the Analamanga time series. Extension of Fig. 3C (main text) depicts the weekly histogram of Ct values from the Analamang timeseries in green, and the resulting Ct distributions at each timepoint from fitted Gaussian process (red) and SEIR (purple) models. Thick line gives the median of distribution for each fit and translucent shading corresponds to the 95% quantiles.

**Fig. S9.**
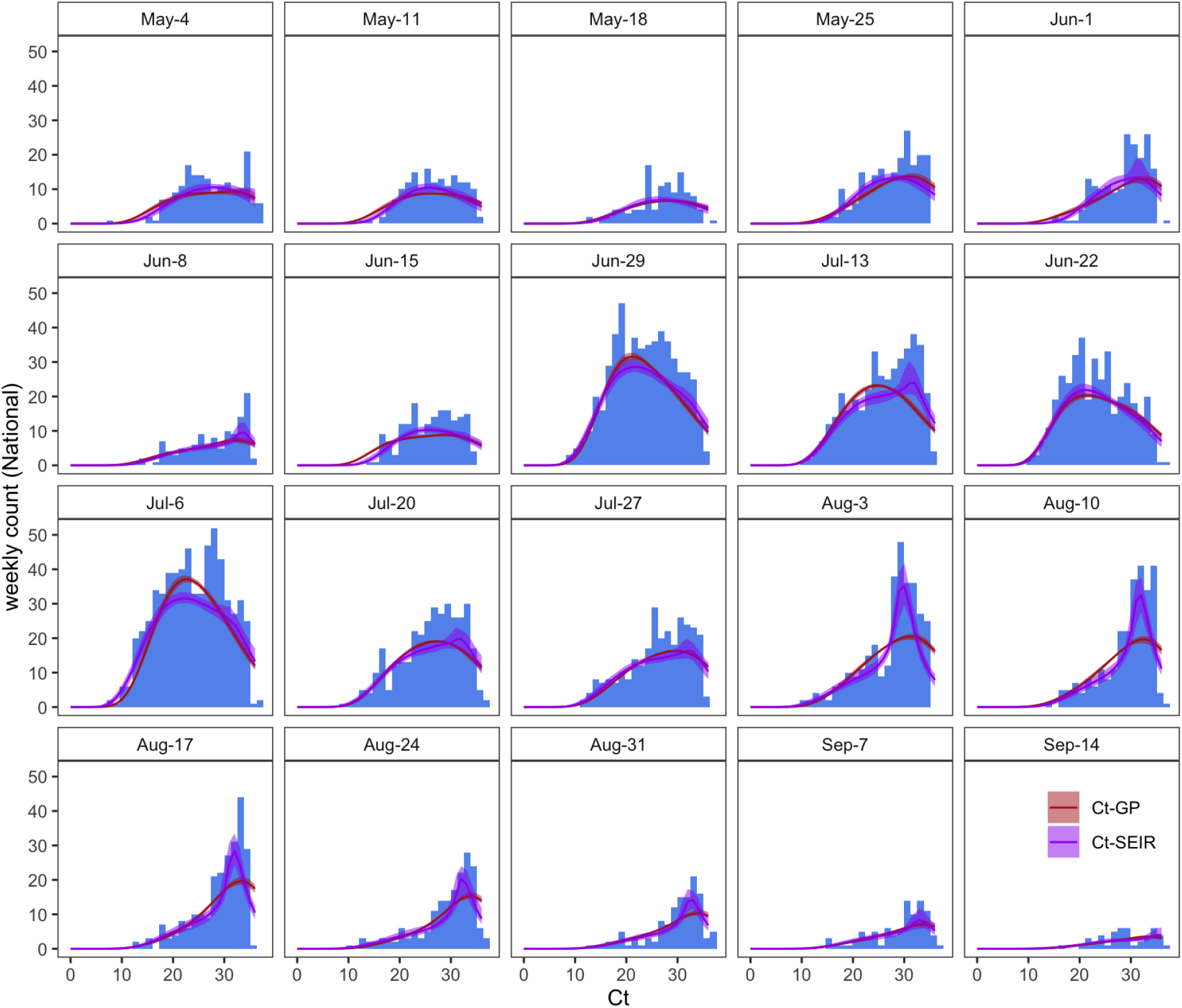
Gaussian process and SEIR Ct-model fits to cross-sectional histograms of Ct values across the Atsinanana time series. Extension of Fig. 3C (main text) depicts the weekly histogram of Ct values from the Atsinanana timeseries in red, and the resulting Ct distributions at each timepoint from fitted Gaussian process (red) and SEIR (purple) models. Thick line gives the median of distribution for each fit and translucent shading corresponds to the 95% quantiles.

## Supplementary Table Captions

**Table S1. Model parameters (for viral kinetics + SEIR + Gaussian process models)**. Fixed and estimated parameters and corresponding descriptions for all parameters used in viral kinetics, SEIR, and Gaussian process models fit to cross-sectional C_t_ distributions across all three regions.

**Table S2. Dilution series of C**_**t**_ **value returned from disparate RT-qPCR platforms inoculated with three virus isolates**. Raw data from tissue culture inoculations, showing C_t_ value resulting from RNA extracted after virus isolate inoculations in cell culture. Data are organized by the test and target used to assay each replicate of each isolate across the dilution series.

**Table S3. Fitted linear mixed effect regression models to tissue culture inoculations in Table S2**. Slopes and y-intercepts of linear mixed effects regression models fit to tissue culture dilutions series in Table S2. Regression lines are visualized in Fig. S1; note that the predictor variable of “dilution”, a proxy for viral load, is modeled on a log10 scale.

**Table S4. Summary output from longitudinal Ct GAMs (Fig. 2D) by region**. Summary from longitudinal GAMs fit to variation in population-level Ct across all three regions (Atsinanana, A., Analamanga, B., National, C.) by date.

**Table S5. Summary output from individual trajectory GAMs**. Summary from individual trajectory GAM used to parameterize within-host viral kinetics model (A.) and individual trajectory GAM used to query the effect of symptom status on Ct variation independent of date of infection onset (B.).

**Table S6. Summary output from population-level symptom status GAM**. Summary from population-level GAM used to query the effect of symptom status on Ct variation across all regions, independent of date.

**Table S7. Convergence diagnostics and posterior quantiles for Gaussian process-Ct models**. Convergence diagnostics, including 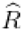, the potential scale reduction factor (values <1.1) and effective population size (values > 200) for all parameters estimated across all three regions via Gaussian process model fit to C_t_ time series. Table also includes mean and 95% posterior quantile for each parameter estimate from the fitted model.

**Table S8. Convergence diagnostics and posterior quantiles for SEIR-Ct models**. Convergence diagnostics, including 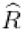, the potential scale reduction factor (values <1.1) and effective population size (values > 200) for all parameters estimated across all three regions at all timepoints via mechanistic SEIR model fit to C_t_ time series. Table also includes mean and 95% posterior quantile for each parameter estimate from the fitted model.

